# High Rates of Rapid Antigen Test Positivity After 5 days of Isolation for COVID-19

**DOI:** 10.1101/2022.02.01.22269931

**Authors:** Emily Landon, Allison H. Bartlett, Rachel Marrs, Caroline Guenette, Stephen G Weber, Michael J. Mina

**Affiliations:** Section of Infectious Diseases & Global Health, Department of Medicine, University of Chicago, Chicago, IL; Infection Prevention & Control Program, University of Chicago Medicine, Chicago, IL; Center for Healthcare Delivery and Innovation, University of Chicago Medicine, Chicago, IL; Section of Pediatric Infectious Diseases, Department of Pediatrics, University of Chicago, Chicago, IL; Office of Occupational Medicine, University of Chicago Medicine, Chicago, IL; Emed, Miami, FL

## Abstract

**Background:** The emergence of the highly transmissible COVID-19 variant, omicron, has resulted in high numbers of breakthrough infections, including among healthcare workers (HCW). Recent CDC recommendations now allow healthcare workers to return to work after day 5 if symptoms have improved, without a requirement for a negative rapid antigen test (RAT).

**Methods:** Fully vaccinated and non-immunocompromised HCW at a large, urban, academic medical center who tested positive for COVID-19 starting in late December, 2021 (when omicron was the predominant circulating strain) were allowed to return to work early if all symptoms had resolved excepting mild, intermittent cough and/or lingering loss of taste/smell, provided a rapid antigen test was negative upon return. Those with negative tests were allowed to return to work with the stipulations that they wear an N95 at all times and take breaks and eat meals apart from others. Those with positive tests on first attempt could return 24-48 hours later to test again for as many days as needed to achieve a negative result or until they completed 10 days of restriction from work.

**Results:** Between January 2, 2022 and January 12, 2022 there were 309 total RAT done on 260 separate HCW on day 5-10 of illness. Overall, 43% (134 of 309) of all RAT were positive between days 5-10. The greatest percent positive RAT was noted among HCW returning for their first test on day 6 (58%). The rate of positivity was greatest (58%) among HCW returning for their first test on day 6. HCW returning on day 8 and 9 were less likely to have a positive test (26%, 19/74). In RAT positive HCW returning for their first test on days 5 or 6 (and for which line intensity was recorded) 49% (25/51) were recorded as having the darkest intensity on their RAT. HCW who test positive on their first test most often remained positive on their second test, with 56% of second tests, aggregated across all days 6-10, remaining positive. Over all first tests performed on days 5-10, boosted HCW were nearly twice as likely to test RAT positive: 53% (75 out of 141) of boosted HCW tested positive.

**Discussion:** More than 40% of vaccinated HCW who felt well enough to work still had positive RAT tests when presenting for a first test between days 5 and 10. Boosted individuals were nearly 3x as likely to result positive on day 5, their first day eligible for return, and ∼2x as likely to result positive on first RAT overall. New guidance provides clearance to exit isolation after 5 days from symptom onset, without the need for a negative rapid antigen test to exit, as long as symptoms are beginning to resolve. Per CDC, the guidance was driven by prior studies, mostly collected before Omicron and before most people were vaccinated or infected, that reported on symptom onset beginning one or more days after peak virus loads. In such an instance, where isolation based on symptom onset often did not begin until peak virus load was already attained, then release from isolation at 5 days would be appropriate. However, reports showing much earlier onset of symptoms, coupled with our own results here demonstrate that the relationship between symptom onset and peak virus load has changed, and 5 days from symptom onset may no longer be an appropriate window to end isolation without a negative rapid antigen test to support safe exit.

**Conclusion:** These results indicate that a substantial proportion of individuals with COVID-19 are likely still contagious after day 5 of illness regardless of symptom status. Early liberation from isolation should be undertaken only with the understanding that inclusion of individuals on day 6-10 of illness in community or work settings may increase the risk of COVID-19 spread to others which, in turn, may undermine the intended benefits to staffing by resulting in more sick workers.

## Background

The emergence of the highly transmissible COVID-19 variant, omicron, has resulted in high numbers of breakthrough infections, including among healthcare workers (HCW). For HCWs, short staffing and patient surges have resulted in a need to return asymptomatic individuals back to the workplace earlier than the previously recommended 10 days after mild COVID-19 infection.

Early studies of COVID-19 viral dynamics and epidemiology indicated that viral shedding and infectiousness began, on average, 2-3 days prior to symptom onset and that maximal virus shedding often began 1 day before symptom onset, with high transmissibility continuing, on average, through the third day of illness.^1^

Based on these findings from prior variants, recent CDC recommendations now allow healthcare workers to return to work after day 5 if symptoms have improved, without a requirement for a negative rapid antigen test (RAT). Updated recommendations acknowledge RAT but do not specifically recommend their use.

Unfortunately, relatively less is known about the relationship between symptom onset and viral dynamics, particularly in vaccinated individuals, infected with the newer omicron variant. Confirming clearance of infectious virus would be ideal prior to returning healthcare workers (HCW) to care for vulnerable individuals. PCR tests often remain positive long after there is no cultivable virus present, making PCR an unsuitable surrogate for clearance to exit isolation^2,3,4,5^. Numerous studies have indicated that Rapid Antigen Tests have superior concordance with cultivable virus, indicating infectious virus, and excellent sensitivity in detecting presence of COVID-19 virus when concomitant PCR tests are positive with a cycle threshold value under 30, which also correlates with presence of cultivable virus^4,5,6^. In addition, numerous studies have demonstrated that darker or more intense positive sample lines on RAT indicate higher amounts of cultivable or infectious virus.^7^

Here, we describe the rate of RAT positivity among HCWs returning to work from isolation in a university hospital setting. We use a semi-quantitative measure of RAT positivity (RAT line intensity) to infer high versus low virus load, corresponding with high versus low infectivity. We discuss the findings in light of the recent CDC guidance for return to work at five days without requirement for a negative Rapid Antigen Test.

## Methods

Fully vaccinated (having received at least 2 doses of a mRNA vaccine) and non-immunocompromised HCW at a large, urban, academic medical center who tested positive for COVID-19 starting in late December, 2021 (when omicron was the predominant circulating strain) were allowed to return to work early if all symptoms had resolved excepting mild, intermittent cough and/or lingering loss of taste/smell, provided a rapid antigen test was negative upon return. Quidell Quickview (San Diego, CA, USA) point of care rapid antigen tests were performed on qualifying HCW by trained hospital staff starting as soon as day 5 of illness (counting from start of symptoms or test date if asymptomatic). Those with negative tests were allowed to return to work with the stipulations that they wear an N95 at all times and take breaks and eat meals apart from others. Those with positive tests on first attempt could return 24-48 hours later to test again for as many days as needed to achieve a negative result or until they completed 10 days of restriction from work. All were allowed to return to work on day 11 without a test, provided symptoms had resolved. For positive tests, intensity of the line was recorded as 1 (more intense than the control line), 2 (same as control line), or 3 (less intense than the control line), in accordance with the instructions for use included by the manufacturer. Due to the urgency for which data is required during this pandemic, only basic analyses of these results are presented here. This evaluation was exempted from requiring IRB approval as Quality Improvement. HCW were offered the opportunity to submit an additional swab for COVID-19 PCR testing as Quality Improvement. Pending IRB approval, these swabs will also undergo virus culture and a more comprehensive evaluation of this data will be performed.

## Results

Between January 2, 2022 and January 12, 2022 there were 309 total RAT done on 260 separate HCW on day 5-10 of illness.

### Day of first return

HCW could present for their first test after isolation anytime between 5 and 10 days of illness as long as symptoms were resolved. The most common day for HCW to return for their first test was day 7 (26%) followed by days 6, 5, 8, 9 and 10, in that order (Table 1, Figure 1). Among HCW receiving 3 doses of mRNA vaccine (boosted), there was little difference in rates of first return between days 5, 6, and 7 (23% - 25%). On the other hand, those receiving only 2 doses mRNA vaccine (unboosted) were most likely to return for their first test on day 7 (28%) compared to the next most common days 8, 6 and 5 (19%, 18%, 16% respectively) (Figure 2).

**Table 1:** RAT Positivity on First Test by Day of Illness and Booster Status.

| Day of Illness |  | Day 5 | Day 6 | Day 7 | Day 8 | Day 9 | Day 10 | Total |
| --- | --- | --- | --- | --- | --- | --- | --- | --- |
| **Total 1<sup>st</sup> Tests** | **n** | **52** | **53** | **68** | **46** | **28** | **13** | **260** |
|  | **% Positive** | **46%** | **58%** | **38%** | **26%** | **25%** | **54%** | **41%** |
| **Not Boosted** | **n** | 19 | 21 | 33 | 23 | 15 | 8 | 119 |
|  | % Positive | 21% | 38% | 24% | 22% | 20% | 50% | 27% |
| **Boosted** | **n** | 33 | 32 | 35 | 23 | 13 | 5 | 141 |
|  | % Positive | 61% | 72% | 51% | 30% | 31% | 60% | 53% |

**Figure 1.**
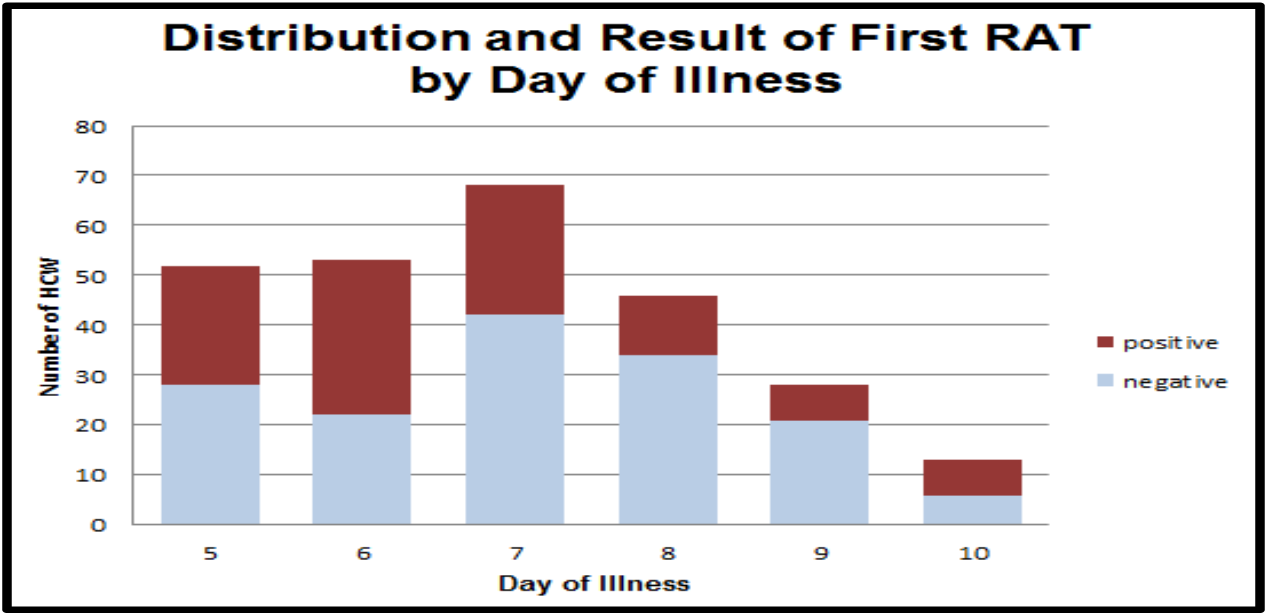

**Figure 2.**
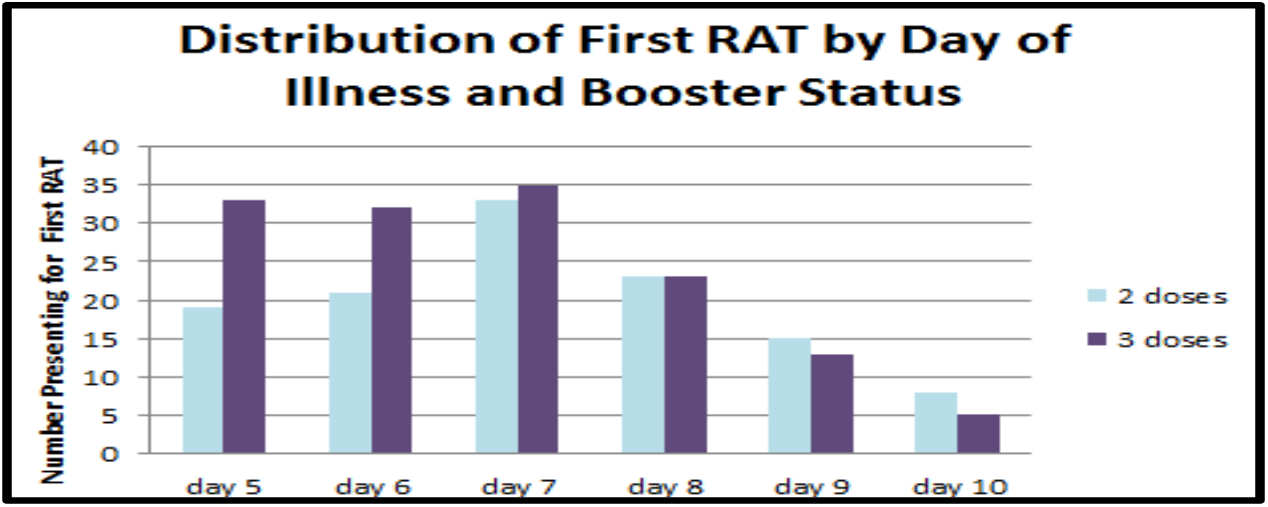

### Rapid Antigen Test positivity

Overall, 43% (134 of 309) of all RAT were positive between days 5-10. The percent positive RAT on the first test performed, regardless of day of first return, was 41% (107 of 260). Among HCW returning on day 5, the percent RAT positive was 46%. The greatest percent positive RAT was noted among HCW returning for their first test on day 6 (58%). After day 6, there was a trend towards decreasing percent positivity upon first test, except for day 10, the last day to receive a first test for return, when 54% of tests were positive (see Table 1).

In RAT positive HCW returning for their first test on days 5 or 6 (and for which line intensity was recorded) 49% (25/51) were recorded as having the darkest intensity on their RAT (Figure 3). Day 6 saw the highest fraction with a very dark positive line, and by day 7 the predominant intensity was medium, followed by day 8 where the predominant intensity was light.

**Figure 3.**
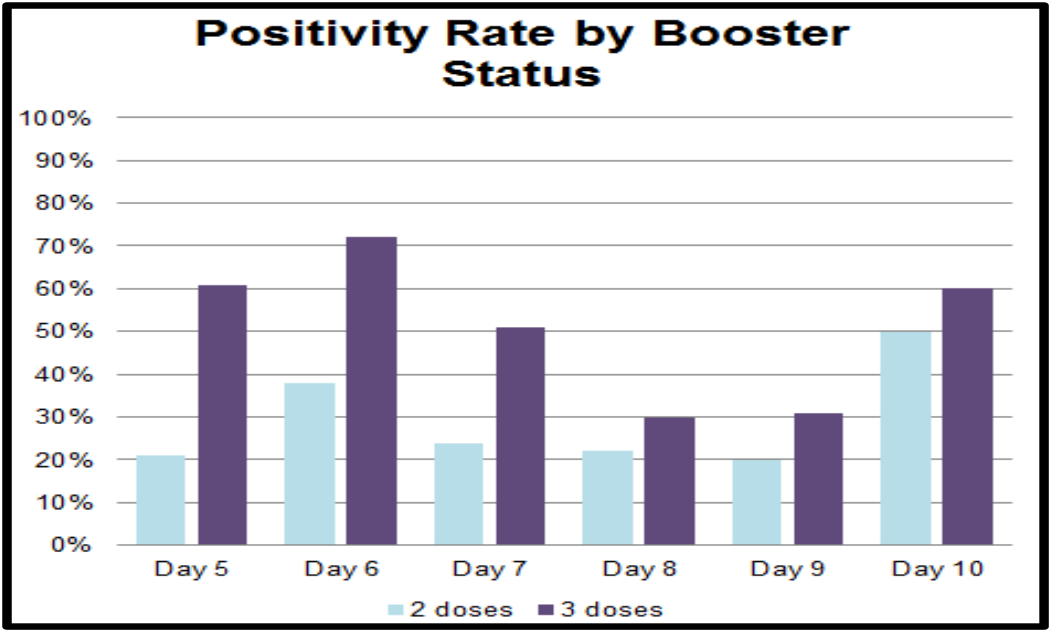

**Figure 4.**
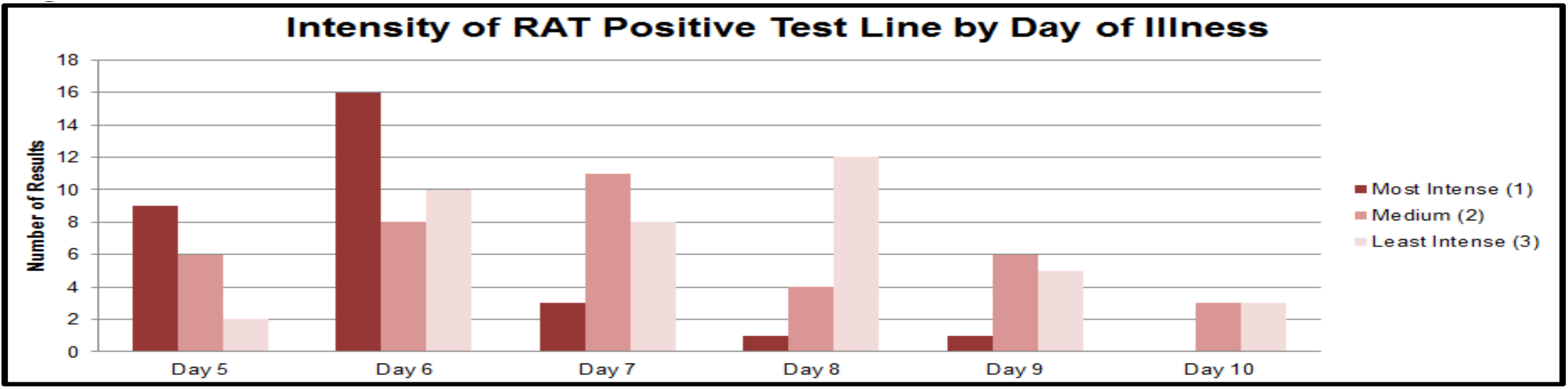

HCW who test positive on their first test most often remained positive on their second test, with 56% of second tests, aggregated across all days 6-10, remaining positive (Table 2).

**Table 2:** Percent positive by test number and day of illness

| **Day of Illness** | **test 1** | **test 2** | **test 3** |
| --- | --- | --- | --- |
| Day 5 | **46%** (24 of 52) | NA | NA |
| Day 6 | **58%** (31 of 53) | **71%** (5 of 7) | NA |
| Day 7 | **38%** (26 of 68) | **33%** (3 of 9) | **50%** (1 of 2) |
| Day 8 | **26%** (12 of 46) | **73%** (8 of 11) | **0%** (0 of 1) |
| Day 9 | **25%** (7 of 28) | **57%** (4 of 7) | **67%** (2 of 3) |
| Day 10 | **54%** (7 of 13) | **0%** (0 of 2) | NA |
| Total | **41%** (107 of 260) | **56%** (20 of 36) | **50%** (3 of 6) |

### Impact of Booster

Crucially, the percent first RAT positive was not uniformly distributed by booster status. Among individuals returning on day 5 for their first test, boosted HCW were nearly 3 times more likely than unboosted HCW to present with a positive RAT (61% positivity among boosted vs. 21% among unboosted). (Table 1 and Figure 2). Over all first tests performed on days 5-10, boosted HCW were nearly twice as likely to test RAT positive: 53% (75 out of 141) of boosted HCW tested positive compared to 27% (32 of 119) for unboosted HCW.

## Discussion

More than 40% of vaccinated HCW who felt well enough to work still had positive RAT tests when presenting for a first test between days 5 and 10. The rate of positivity was greatest (58%) among HCW returning for their first test on day 6. HCW returning on day 8 and 9 were less likely to have a positive test (26%, 19/74).

Individuals returning for their first test on day 10 had a higher rate of positivity, though there were too few HCW in this group (13) to make any conclusions about these results.

Boosted individuals were nearly 3x as likely to result positive on day 5, their first day eligible for return, and ∼2x as likely to result positive on first RAT overall. The increased rate of positivity among boosted individuals persisted across all days. It is plausible to consider that boosted individuals may become symptomatic earlier in the course of illness as a result of faster immune detection of the virus with immune mediated symptom onset. This fits with a notable shift in symptoms observed with “Omicron” towards those indicative of immune activation (coryza, rhinorrhea, cough and sore throat)^8^ versus symptoms noted in early studies done in a completely immunologically naïve population, where symptoms began only after peak virus load was achieved and were most indicative of severe virus mediated damage (most notably loss of smell and difficulty breathing).^9,10^

If virus kinetics in the nasal passage remain largely unperturbed by pre-existing immunity, as has been demonstrated in longitudinal studies of Omicron^11^, then this would manifest as symptom onset days before peak viral load, rather than symptom onset after peak virus load is attained. In such a situation, individuals might then be at or remain near their peak of virus load and infectivity at day 5 from symptom onset. Such an immune mediated phenomenon, driven by a swift anamnestic response, would be most likely to impact recently boosted individuals, followed by vaccinated and previously infected individuals, whereas entirely immunologically naïve individuals would still develop onset of symptoms well into the viruses growth, if not the decay phase, as was the case in almost all data sets collected earlier in the pandemic.^12,13^

This has implications for recent guidance from the the US Centers for Disease Control and Prevention. New guidance provides clearance to exit isolation after 5 days from symptom onset, without the need for a negative rapid antigen test to exit, as long as symptoms are beginning to resolve. Per CDC, the guidance was driven by prior studies, mostly collected before Omicron and before most people were vaccinated or infected, that reported on symptom onset beginning one or more days after peak virus loads. In such an instance, where isolation based on symptom onset often did not begin until peak virus load was already attained then release from isolation at 5 days would be appropriate. This has been noted in numerous studies from earlier in the pandemic demonstrating no cultivable virus at 5 days. However, reports showing much earlier onset of symptoms, coupled with our own results here demonstrate that the relationship between symptom onset and peak virus load has changed, and 5 days from symptom onset may no longer be an appropriate window to end isolation without a negative rapid antigen test to support safe exit.

The intensity of the line on RAT correlates with the level of virus in the sample^7,14^. A dark line indicates a strong positive with a high level of virus and is usually seen when people are at or near peak virus load. A light or faint line indicates a lower level of virus and is usually associated with lower but not absent amounts of cultivable virus that can still represent infectious virus. These data, demonstrate a high fraction of very dark high intensity RAT lines at day five and suggests that the relationship between symptom onset and high virus titers may have shifted. Given the higher fraction positive among recently boosted individuals, putatively, this may be a result of earlier immunological activation and onset of immune mediated symptoms. Our data strongly suggests that safely exiting isolation as early as day 5 since symptom onset should include a negative RAT prior to exit, regardless of vaccination or booster status and should certainly not be predicated on symptom resolution alone.

Further analyses of cultivable virus in samples will help to refine our data. While most studies looking at the relationship between cultivable (i.e. infectious) virus and RAT positivity have defined strong concordance between the two, it may be the case, for example, that after a certain number of days since symptom onset a very faint line (but unlikely a dark or medium line) on a RAT may no longer indicate highly cultivable virus that is infectious to others.

## Limitations

Because participation in this program was voluntary and limited to work areas with critical staffing needs, not every HCW returned every day for testing. Additionally, we did not collect data about when HCW’s symptoms resolved with respect to their presentation for testing and we believe many non-medical factors may have influenced the timing of the RAT including date of next scheduled shift and/or development of a new critical staffing need in their work area. This is perhaps most notable by the increased fraction positive on day 10, the last day individuals were asked to isolate before returning to work without a negative test.

Because RAT correlates well with PCR positivity with cycle threshold below 30 and that, in turn, correlates with recovery of cultivable virus, it is reasonable to believe that HCW with positive RAT are likely contagious.

However, this has not been established for omicron. This team has collected over 150 samples for corroboratory culture and PCR testing as of January 15 and will update this paper with those results when available.

## Conclusion

Despite the limitations noted, these results indicate that a substantial proportion of individuals with COVID-19 are likely still contagious after day 5 of illness regardless of symptom status. Early liberation from isolation should be undertaken only with the understanding that inclusion of individuals on day 6-10 of illness in community or work settings may increase the risk of COVID-19 spread to others which, in turn, may undermine the intended benefits to staffing by resulting in more sick workers. RAT testing can identify those at lower risk of transmitting COVID and should be employed alongside strict adherence to masking whenever vulnerable individuals are present including the unvaccinated, elderly, and/or immunocompromised.

## Data Availability

Deidentified data produced in the present study are available upon reasonable request to the authors.

## References

1. Lau EHY, Leung GM. Reply to: Is presymptomatic spread a major contributor to COVID-19 transmission? Nature Medicine 2020;26:1534–1535. https://www.nature.com/articles/s41591-020-1049-3

2. Mina MJ, Parker R, Larremore DB. Rethinking Covid-19 Test Sensitvity—A Strategy for Containment. NEJM. 2020;383;22 e120–123. https://www.nejm.org/doi/full/10.1056/nejmp2025631

3. Tom MR, Mina MJ. To Interpret the SARS-CoV-2 Test, Consider the Cycle Threshold Value. Clin Infect Dis 2020;71 2252–4. https://academic.oup.com/cid/article/71/16/2252/5841456?login=true

4. Pekosc A, Parvu V, Li M, et al. Antigen-based Testing but Not Real-Time Polymerase Chain Reaction Correlates With Severe Acute Respiratory Syndrome Coronavirus 2 Viral Culture. Clin Infect Dis 2021;73 e2861–6. https://academic.oup.com/cid/article/73/9/e2861/6105729

5. Parvu V, Gery DS, Mann J, et al. Factors the Influence the reported Sensitivity of Rapid Antigen Testing for SARS-CoV-2. Front Microbiol 12:714242. Doi:10.3389/fmicb.2021.714242 https://www.frontiersin.org/articles/10.3389/fmicb.2021.714242/full

6. Schrom J, Marquez C Pilarowski, et al. Direct Comparison of SARS CoV-2 Nasal RT-PCR and Rapid Antigen Test (BinaxNow) at a Community Testing Site During an Omicron Surge MedRxiv Jan 10, 2021. https://www.medrxiv.org/content/10.1101/2022.01.08.22268954v2

7. Toptan T, Eckermann L, Pfeiffer, et al. Evaluation of a SARS-CoV-2 rapid antigen test: Potential to help reduce community spread? J Clin Virol. 2021;135:104713 https://www.sciencedirect.com/science/article/pii/S1386653220304558?via%3Dihub

8. Newton AH, Cardani A, Braciale TJ. The host immune response in respiratory virus infection: balancing virus clearance and immunopathology. Semin Immunopathol. 2016. 38:471–482. https://www.ncbi.nlm.nih.gov/pmc/articles/PMC4896975/pdf/281_2016_Article_558.pdf

9. Gengler I, Wang JC, Speth MM, Sedaghat AR. Sinonasal pathophysiology of SARS-CoV-2 and COVID-9: A systematic review of the current evidence. Laryngoscope Investigative Otolaryngology. 2020;5:354–9. https://doi.org/10.1002/lio2.384

10. Subbarao K, Mahanty S. Respiratory Virus Infections: Understanding COVID-19. Immunity. 2020;52:905–9. https://doi.org/10.1016/j.immuni.2020.05.004

11. Hay J, Kissler S, Fauver JR, Mack C, Tai CG. Viral dynamics and duration of PCR positivity of the SARS-CoV-2 Omicron Variant. Pre-Print https://dash.harvard.edu/bitstream/handle/1/37370587/omicron_ct.1-13-22.4.pdf?sequence=1&isAllowed=y

12. Singanayagam A, Patel M, Charlett A et al. Duration of infectiousness and correlation with RT-PCR cycle threshold values in cases of COVID-19, England, January to May 2020. Euro Surveill. 2020;25(32):pii=2001483. https://doi.org/10.2807/1560-7917.ES.2020.25.32.2001483

13. Byrne AW, McEvoy D, Collins AB, et al. Inferred duration of infectious period of SARS-CoV-2: rapid scoping review and analysis of available evidence for asymptomatic and symptomatic COVID-19 cases. BMJ Open 2020;10:e039856 doi:10.1136/bmjopen-2020-039856

14. Pilarowski G, Lebel P, Sunshine S, et al. Performance characteristics of a rapid severe acute respiratory syndrome coronavirus 2 antigen detection assay at a public plaza testing site in San Francisco. Journ Infect Dis 2021;223:1139–44. https://academic.oup.com/jid/article/223/7/1139/6061974

